# Clinical-grade Cuffless Blood Pressure Monitoring via Deep-tissue Diffuse Speckle Pulsatile Flowmetry

**DOI:** 10.64898/2026.06.19.26356074

**Authors:** Tristan Wen Jie Choo, Alicia Ann May Yap, Ariffin Kawaja, Victor T. T. Chao, Chirk Jenn Ng, Malini Olivo, Renzhe Bi

## Abstract

Blood pressure (BP) is a vital sign which is measured to diagnose and manage hypertension. However, current methods to measure BP use inflatable cuffs which cause discomfort and limit the frequency at which measurements can be made, or intra-arterial catheters which are invasive and pose infection risks. Here, we propose and evaluate the use of Diffuse Speckle Pulsatile Flowmetry (DSPF) as a cuffless BP measurement method to address these limitations. DSPF is a laser speckle-based technique which simultaneously records blood flow rate and blood volume (i.e. photoplethysmography or PPG) signals from relatively deep vascular tissue. Using information from these signals, we studied DSPF’s effectiveness in measuring systolic BP (SBP) and diastolic BP (DBP) through an outpatient study in which 133 patients were recruited, and in measuring beat-to-beat BP waveforms through an inpatient study in which two patients were recruited. In the outpatient study, the DSPF method was able to achieve mean absolute errors (MAEs) of 4.17 mmHg and 2.42 mmHg for SBP and DBP respectively compared to conventional cuff-based methods. It was also able to fulfil the requirements of the AAMI/ESH/ISO 81060-2:2018 standard for BP measurement devices and attain an ‘A’ grade according to the British Hypertension Society’s grading scheme. For the inpatient study, it produced BP waveforms which had MAEs of 2.35 mmHg and 3.06 mmHg compared to arterial-line measurements for the two patients, respectively. Compared to PPG which has been studied more extensively as a cuffless BP measurement method, we found through ablation studies that DSPF was able to reach significantly lower MAEs and hence better accuracies. DSPF augments the performance of PPG-only methods by leveraging additional information from the blood flow rate signal, and we therefore find it to be a superior cuffless BP measurement method which can potentially be used in outpatient, inpatient, and remote settings.

## Introduction

Regular and frequent blood pressure (BP) measurements are critical for the management of cardiovascular diseases such as hypertension^1,2^. At present, however, the devices and techniques which are conventionally used to measure BP in clinical and remote settings have several limitations; cuff-based sphygmomanometers are unable to make continuous measurements and cause discomfort to patients^3,4^, while intra-arterial catheters are invasive and pose infection risks^5^. Although photoplethysmography (PPG) sensors have more recently been introduced to measure BP indirectly from blood volume measurements^6–10^, their use relies on information from relatively superficial blood vessels in the microvascular bed which is potentially problematic^11,12^.

To overcome these limitations, we propose the use of Diffuse Speckle Pulsatile Flowmetry (DSPF) to make continuous, non-invasive, and cuffless BP measurements. DSPF, alternatively known as speckle contrast optical spectroscopy, is an optical technique which makes simultaneous measurements of blood flow rate and blood volume (i.e. PPG) over time in relatively deep blood vessels (up to a depth of approximately 15 mm). To date, it has been demonstrated to measure perfusion in diabetic feet^13^, assess endothelial function^14^, and observe the vascular response to cold shock^15^. By leveraging the biomechanical links between flow, volume, and pressure, BP can also be modelled and measured from the information in DSPF signals. This paper presents results from two clinical studies to validate DSPF’s effectiveness in achieving this in outpatient and inpatient settings.

Initial investigations to evaluate the feasibility of DSPF in measuring BP have shown early signs of promise by fulfilling criterion 1 of the Association for the Advancement of Medical Instrumentation/European Society of Hypertension/International Organization for Standardisation’s (AAMI/ESH/ISO) standard (ISO 81060-2:2018) for the validation of BP measuring devices^16^. These studies reported average measurement errors under the threshold of 5 mmHg as well as a standard deviation of errors under the requirement of 8 mmHg. In addition, they showed that features from the blood flow waveform significantly improved the measurement of BP^17,18^. However, these studies only involved healthy subjects with normotensive BP values at rest, rather than baseline hypertensive and hypotensive individuals. Moreover, the number of subjects did not exceed 30, a sample size considerably lower than the minimum requirement of 85 as stipulated by the AAMI/ESH/ISO standard. To build on these early results, we aim to validate the effectiveness of DSPF in a larger clinical population presenting a wider range of BP values to achieve closer adherence to standard’s requirements, with the aim of progressing towards clinical translation.

Toward this end, we conducted an outpatient study in Eunos Polyclinic, a primary care facility in Singapore where 133 patients with different demographic profiles and presenting with a wide range of systolic BPs (SBPs) and diastolic BPs (DBPs) were recruited (Table 1). An intentional effort was made to ensure that ≥ 30% of the recruited patients were male and ≥ 30% were female; ≥ 5% of the reference SBP readings from the sample were ≤ 100 mmHg, ≥ 5% were ≥ 160 mmHg, and ≥ 20% were ≥ 140 mmHg; while ≥ 5% of the reference DBP readings were ≤ 60 mmHg, ≥ 5% were ≥ 100 mmHg, and ≥ 20% were ≥ 85 mmHg. This was done in accordance with the requirements of AAMI/ESH/ISO 81060-2:2018^16^. Cuff-based BP measurements as well as DSPF signals from the thumb, wrist, and elbow were obtained from these patients (Fig. 1a). In addition, we also conducted an inpatient study at a specialised cardiology centre, the National Heart Centre Singapore, to validate DSPF’s effectiveness in making continuous measurements of the BP waveform from beat to beat. Two warded patients were recruited for this study, each having an intra-arterial catheter inserted in their radial artery and DSPF sensors at the thumb and elbow (Fig. 1b). To the best of our knowledge, this is the first study of its kind to be reported.

**Fig. 1:**
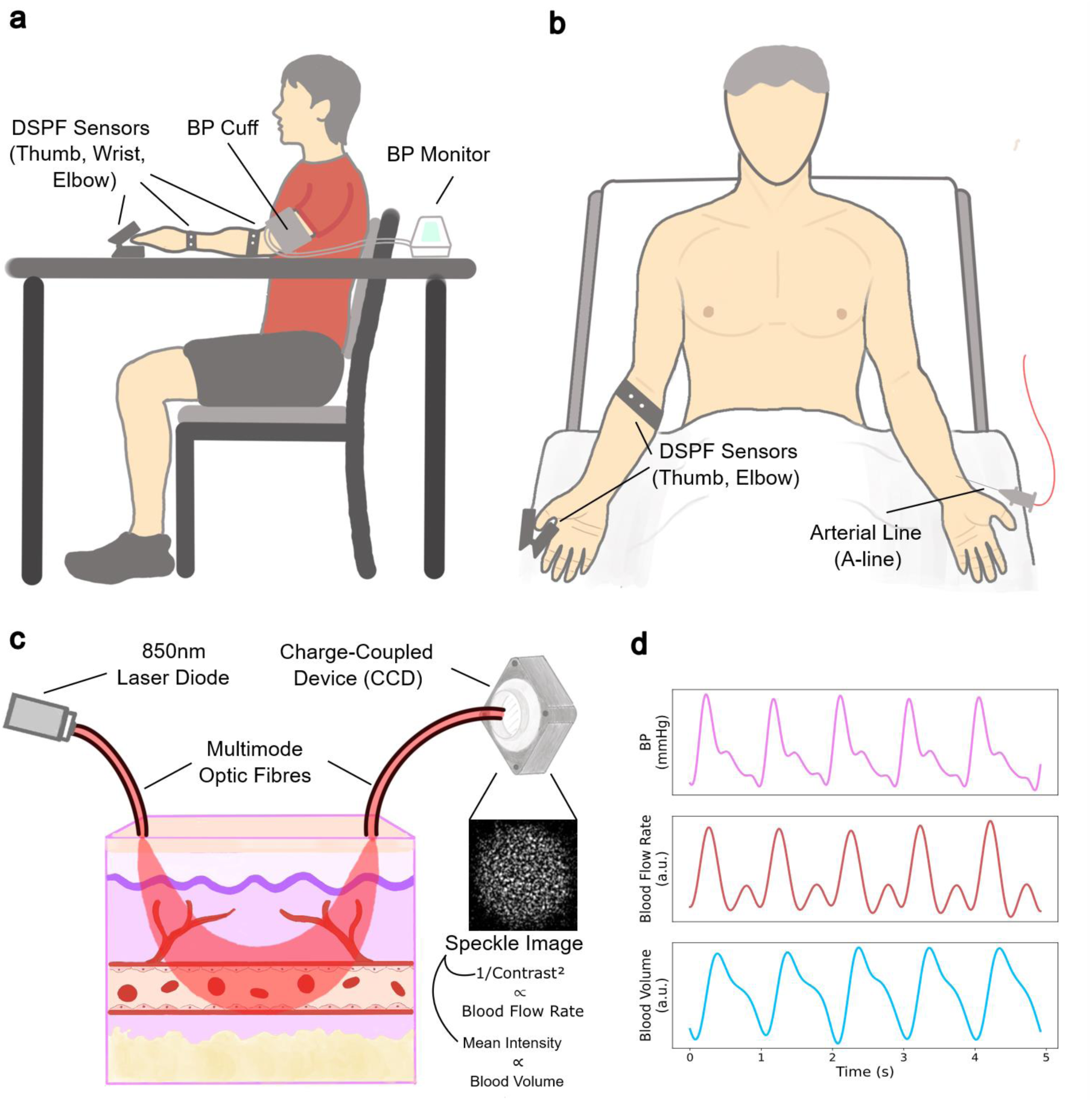
The Diffuse Speckle Pulsatile Flowmetry (DSPF) system for BP monitoring in outpatient and inpatient settings. a) Experimental setup for the outpatient study. DSPF sensors were placed at the thumb, wrist, and elbow of each participating subject. Blood flow rate and blood volume signals were collected from these anatomical locations and used to determine SBP and DBP. Reference BP values were obtained from a conventional sphygmomanometer. b) Experimental setup for the inpatient study. Each subject had DSPF sensors placed over the thumb and elbow, as well as an arterial line (A-line) inserted into the radial artery of the contralateral arm. Blood flow and blood volume waveforms from the DSPF sensors were used to construct the BP waveform for which the reference was provided by the A-line. c) Schematic of the operating principle of a set of DSPF sensors. A 850 nm laser diode shines coherent light into a ‘source’ multimode fibre which contacts the skin, following which the emitted photons are backscattered by red blood cells in relatively deep arteries and arterioles. The backscattered photons are collected by another multimode ‘detector’ fibre which transmits them to a charge-coupled device to generate a laser speckle pattern. Statistical measures from the speckle pattern are then used to determine instantaneous blood flow rate and blood volume; the inverse of the speckle contrast squared is proportional to the blood flow rate while the mean pixel intensity is proportional to the blood volume. d) Representative waveforms of BP from the A-line (top), blood flow rate (middle), and blood volume (bottom).

**Table 1:** Demographic characteristics of the subjects in the outpatient study.

| Demographic Characteristic | Group | Number of Subjects | % of sample |
| --- | --- | --- | --- |
| Age | 21-30 | 7 | 5.26 |
|  | 31-40 | 16 | 12.03 |
|  | 41-50 | 17 | 12.78 |
|  | 51-60 | 28 | 21.05 |
|  | 61-70 | 35 | 26.32 |
|  | 71-80 | 23 | 17.29 |
|  | 81-90 | 7 | 5.26 |
| Gender | Male | 66 | 49.62 |
|  | Female | 67 | 50.38 |
| Ethnicity | Chinese | 91 | 68.42 |
|  | Malay | 22 | 16.54 |
|  | Indian | 13 | 9.77 |
|  | Others | 7 | 5.26 |
| SBP | ≤ 80 mmHg | 1 | 0.75 |
|  | 81-100 mmHg | 15 | 11.28 |
|  | 101-120 mmHg | 48 | 36.09 |
|  | 121-140 mmHg | 39 | 29.32 |
|  | 141-160 mmHg | 19 | 14.29 |
|  | 161-180 mmHg | 10 | 7.52 |
|  | > 180 mmHg | 1 | 0.75 |
| DBP | ≤ 60 mmHg | 8 | 6.02 |
|  | 61-70 mmHg | 35 | 26.32 |
|  | 71-80 mmHg | 44 | 33.08 |
|  | 81-90 mmHg | 26 | 19.55 |
|  | 91-100 mmHg | 13 | 9.77 |
|  | 101-110 mmHg | 6 | 4.51 |
|  | > 110 mmHg | 1 | 0.75 |

A DSPF sensor comprises a pair of multimode optic fibres which each have one end in contact with the skin (Fig. 1c). Coherent light from a laser diode passes through one fibre (the ‘source’ fibre) to illuminate the underlying tissue, emitting photons that are subsequently backscattered by red blood cells (RBCs) which represent moving scatterers. These backscattered photons enter and pass through a second fibre (the ‘detector’ fibre) to a charge-coupled device (CCD) which generates a laser speckle pattern. Statistical measures from the speckle pattern can then be calculated to provide information about blood flow rate and blood volume; the square of the speckle pattern’s contrast is inversely proportional to the instantaneous blood flow rate, while the average pixel intensity is proportional to the instantaneous blood volume^19^. By recording the speckle pattern over time, two signals are produced; one which reflects blood flow rate, and another which reflects blood volume (i.e. PPG) (Fig. 1d).

Compared to PPG, DSPF possesses several advantages which provides the rationale for its clinical utility as a non-invasive and cuffless BP measurement device. Firstly, it offers a larger penetration depth which allows for the interrogation of deeper vasculature (i.e. arteries, arterioles) compared to PPG. The limited penetration depth of PPG restricts its use to the monitoring of smaller vessels and the microcirculation which are further downstream in the vascular hierarchy; the hemodynamics within these vessels differ significantly from the hemodynamics in the brachial artery where cuff-based BP measurements are obtained, or the radial artery where arterial line (A-line) measurements are typically made. By utilising the hemodynamic information in the deeper vessels which are more closely linked to the brachial and radial arteries, BP measurements can theoretically be modelled with higher accuracies. Secondly, the DSPF technique observes blood flow patterns in addition to PPG, providing additional information that is valuable for the determination of BP.

To analyse this hypothesised benefit that DSPF provides over PPG, we performed ablation studies to evaluate how using blood flow rate or PPG in isolation or in combination would affect the accuracy of BP measurements. When using both modalities together, we also investigated the relative importance of each modality to identify which was the stronger contributor to the determination of BP. Additionally, we compared the quality of the blood flow rate and PPG waveforms to determine which modality would be more robust for use in practical settings. These collectively represent new characterisations of the DSPF system that further the current understanding of its use in BP measurement and benchmark its performance against PPG which is more commonly studied as a cuffless BP measurement method.

## Results

### Outpatient Study

For the outpatient study, the blood flow rate and blood volume signals obtained from each patient were first pre-processed to identify pulses with acceptable signal quality, before a set of morphological and statistical features (Fig. 2a) was extracted from the acceptable waveforms of each signal (see Methods). These features, such as the maximum blood flow rate and the increase in blood volume during a cardiac cycle, provide information that is useful for characterising the hemodynamic profile of a pulse. As such, they were used to train machine learning models for SBP and DBP prediction, similar to PPG and/or electrocardiography (ECG)-based approaches^20–28^.

**Fig. 2:**
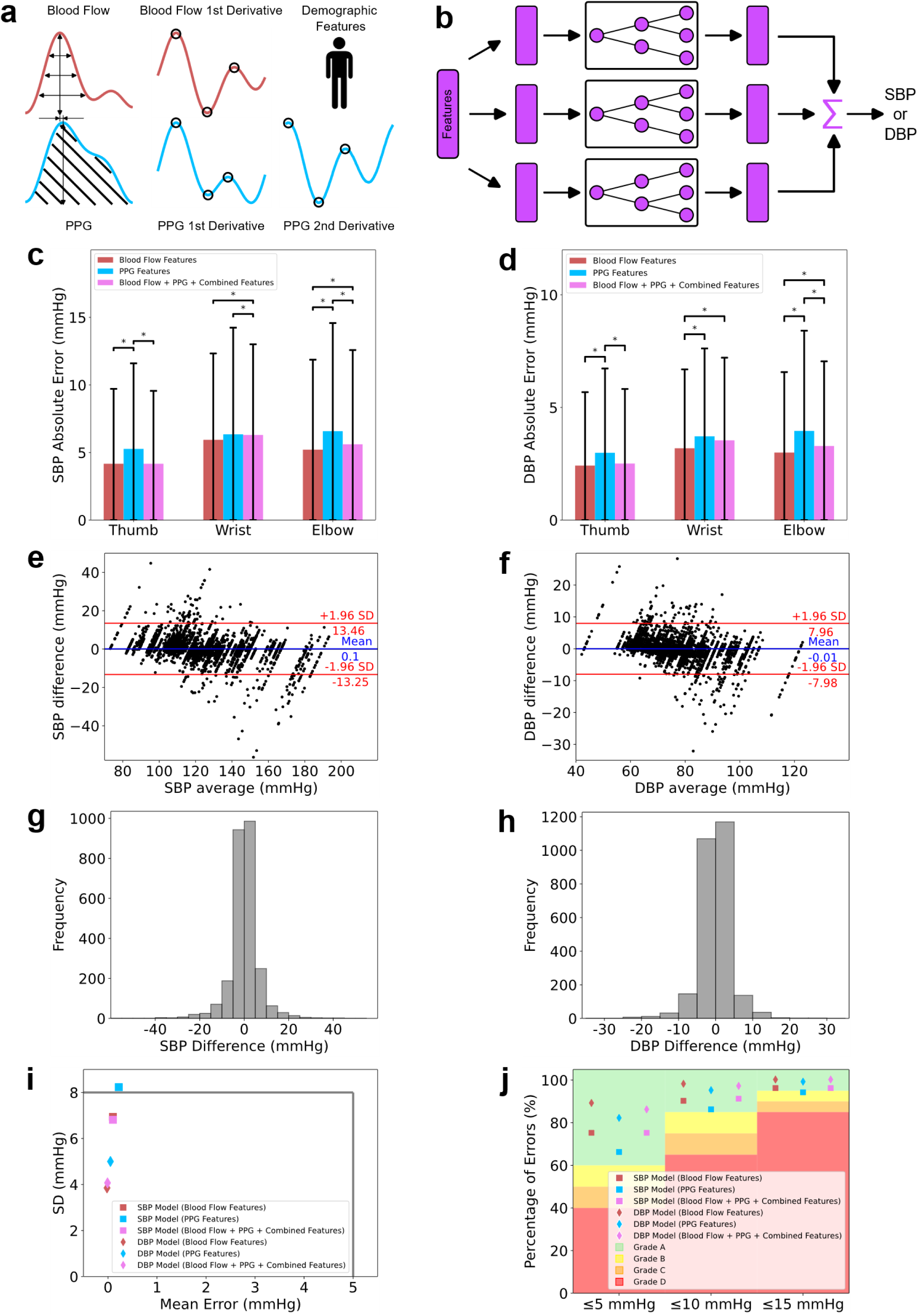
Validation of the DSPF system in the outpatient study. a) Features related to amplitudes, widths, areas, timings and values of fiducial points were extracted from the blood flow and PPG waveforms. Demographic features from each subject were also obtained. b) XGBoost models were trained on different sets of blood flow, PPG, and combined features in an ablation study to estimate SBP or DBP. c-d) Bar charts showing the comparison of MAEs for SBP (c) and DBP (d) across the three different XGBoost models trained on data at each anatomical location where DSPF sensors were placed. Error bars represent standard deviation while asterisks represent statistical significance (p < 0.05) between absolute errors from pairs of XGBoost models. e-f) Bland-Altman plots showing the differences in SBP (e) and DBP (f) between the DSPF-measured and sphygmomanometer-obtained values. Blue lines show the mean difference while red lines show 1.96 standard deviations above and below the mean difference. Good agreement between the two devices is observed through the low mean difference and standard deviations. g-h) Histograms show that the distribution of differences in SBP and DBP as measured by the two devices is approximately normal, indicating that 95% of the differences fall within ±1.96 standard deviations of the mean. i) Benchmarking of the performance of the DSPF system for SBP and DBP measurement against criterion 1 of the AAMI/ESH/ISO 81060-2018 standard shows that using blood flow features or a combination of blood flow and PPG features meets the stipulated requirements whereas using PPG features alone does not. This suggests that information from the DSPF blood flow signal improves the accuracy of PPG-only methods which may not be sufficiently accurate

The blood flow waveforms obtained using DSPF have a similar morphology to those obtained via Doppler ultrasound. Many features from the Doppler ultrasound blood flow waveform, such as the peak systolic speed, end diastolic speed, and resistive index^29^ can be calculated from its DSPF counterpart and were appropriately determined. In addition, we extracted other features inspired by those commonly extracted from PPG waveforms, such as pulse widths and the augmentation index^20,30^. These were considered to bear valuable information for the measurement of BP.

Furthermore, since the derivative of volume with respect to time represents the volumetric flow rate, the blood flow waveform is an analogue to the first derivative of the PPG waveform, while its first derivative can be likened to the second PPG derivative. Several features from the waveform of the second PPG derivative have been found to be strongly associated with BP^31,32^, thus we calculated these features from the first derivative of the blood flow waveform. The full set of features extracted from the blood flow waveform is detailed in Supplementary Table 1.

The waveforms of the blood volume or PPG signal bear the classical systolic peak, dicrotic notch, and diastolic peak which are typically present^33^. To complement the information provided by the features obtained from the blood flow waveform, we extracted commonly used PPG features which also reflect the underlying hemodynamics and biomechanics of the vessels under study^20,34–38^. Features from the first and second derivatives of the PPG waveform which have been found to be useful predictors of BP were also extracted^31,32,38,39^. These PPG-based features can be found in Supplementary Table 2.

An additional advantage of DSPF is that it allows features involving both the blood flow and PPG waveforms to be extracted. The interaction of these two waveforms is potentially of physiological importance as together they can reflect the viscoelasticity (or stiffness) of vascular tissue. One feature of note is the time delay between the systolic peaks of the two waveforms; the systolic peak of the blood flow waveform occurs prior to that of the PPG waveform as the vessel takes time to expand to accommodate the inflow of blood during systole. The duration of this delay is informative, as smaller delays imply that the vascular tissue is stiff and takes a shorter time to reach its full expansion during the cardiac cycle, whereas larger delays indicate that the tissue is more compliant. Features such as these are described in Supplementary Table 3, and we henceforth refer to them as ‘combined features’. Blood flow features, PPG features, combined features, and demographic features from each patient were used to train XGBoost regression models for the estimation of SBP and DBP (Fig. 2b). In some patients, however, there was an insufficient number of acceptable PPG waveforms from which features could be extracted due to noise in the PPG signal. We thus trained models for SBP and DBP estimation which incorporated only blood flow and demographic features to determine the performance of DSPF in the absence of viable PPG waveforms. In addition, we also trained models which only included PPG and demographic features to facilitate a modality-wise comparison between the blood flow and PPG modalities. The development and evaluation of these three types of models effectively represented an ablation study. To ensure uniform computational complexity, the number of features used to train each model was kept constant at 30, with feature selection being implemented by the XGBoost algorithm. Separate models were trained on DSPF data from the thumb, wrist, and elbow with the motivation of achieving BP measurement from a single anatomical site, and different models were trained to measure SBP and DBP.

A five-fold cross-validation was applied for all models, and the mean absolute error (MAE) was used to evaluate each one’s performance (Fig. 2c, d). To determine statistical significance, we performed the Kruskal-Wallis test to identify whether at least one of the three models trained on DSPF data from each anatomical location was performing significantly differently from the other two, and if so, then subsequent pairwise comparisons using Dunn’s test with Bonferroni’s correction to identify which specific model pairs had a statistically significant difference.

The best-performing models were those trained on DSPF data from the thumb; the models trained using blood flow, PPG, and combined features from the thumb had MAEs of 4.17 mmHg for SBP and 2.51 mmHg for DBP, while those trained using blood flow features had MAEs of 4.17 mmHg for SBP and 2.42 mmHg for DBP. Notably, both these types of models significantly outperformed the models trained using PPG features; these had an MAE exceeding 5 mmHg for SBP and an MAE of 2.99 mmHg for DBP.

At the wrist and elbow, the MAEs of the models trained using blood flow features or using all available features were similarly lower than those produced by the models trained using PPG features. However, the models trained using all features had higher MAEs compared to those trained on blood flow features. This can be attributed to the lower number of acceptable pulses which we discuss further below. Nevertheless, all SBP models had MAEs < 7 mmHg while all DBP models had MAEs < 4 mmHg.

Good agreement between the DSPF-derived and cuff-based SBP and DBP values was also observed in Bland-Altman plots (Fig. 2e, f). The mean errors (ME) for the models trained on all features from the blood flow and PPG signals obtained from the thumb were 0.10 ± 7.24 mmHg and -0.01 ± 4.07 mmHg for SBP and DBP, respectively. Histograms of the errors also showed an approximate normal distribution (Fig. 2g, h), suggesting that 95% of all errors fall within ±1.96 standard deviations of the mean (-13.25 to 13.46 mmHg for SBP, -7.98 to 7.96 mmHg for DBP). These results meet criterion 1 of the AAMI/ESH/ISO standard which requires a mean error of < 5 mmHg and a standard deviation of errors < 8 mmHg^16^ (Fig. 2i). In addition, they also meet the ‘A’ grade criteria of the British Hypertension Society’s (BHS) protocol for the evaluation of BP measuring devices, which requires 60% of the absolute errors to be ≤ 5 mmHg, 85% to be ≤ 10 mmHg, and 95% to be ≤ 15 mmHg^40^ (Fig. 2j).

On the contrary, the SBP model trained using PPG features from the thumb did not meet the AAMI/ESH/ISO and BHS criteria. The standard deviation of its errors exceeded 8 mmHg, and it only achieved a ‘B’ grade under the BHS grading system, having just 93% of absolute errors≤ 15 mmHg. This suggests that the information provided by the additional features obtained from the blood flow signal is highly beneficial for meeting the clinical standards applicable to cuffless BP measurement devices, and that using PPG alone may be insufficient for fulfilling these requirements.

### Inpatient Study

While determining SBP and DBP values may be sufficient and more practical in outpatient and remote settings, measuring beat-to-beat BP is often critical for patients who are warded for conditions such as stroke, septic shock, and traumatic brain injury^41–43^. Continuous BP measurements help to alert physicians and nurses to cardiac emergencies, and are useful for observing the effects of vasoactive medications at fine temporal detail. In addition, real-time BP monitoring is vital for perioperative care^44^.

The BP waveform is rich in information, possessing morphological signatures, shapes, and patterns which cannot be observed from isolated SBP or DBP measurements. Such information can reveal hidden pathophysiology and inform diagnoses of complex conditions which are difficult to determine otherwise. To tailor the applicability of the DSPF system to the inpatient setting, we evaluated its ability to produce entire BP waveforms.

The DSPF signals from the two subjects recruited for the inpatient study were pre-processed in a similar manner as in the outpatient study to identify blood flow and PPG waveforms of acceptable quality. Instead of extracting features from individual pulses, however, we trained patient-specific U-Nets to construct the BP waveform using various combinations of blood flow and PPG pulses from the thumb and elbow. The U-Net is a type of neural network architecture which has been effectively used for biomedical image segmentation^45^ and the formation of biosignals such as the ECG^46^, respiration^47^, and BP^48^ signals. They comprise an encoder branch which learns abstractions of data as well as a decoder branch which captures finer detail to produce a mapping between an input and output sequence. Its architecture was therefore well-suited to our task of translating the blood flow and/or PPG waveforms into BP waveforms.

Five U-Nets were trained for each patient in an ablation study format; the first used only blood flow waveforms from the thumb, the second used only PPG waveforms from the thumb, and the third used blood flow waveforms from the elbow. The fourth used a combination of the blood flow and PPG waveforms from the thumb, and the fifth used both types of waveforms from the thumb as well as the blood flow waveforms from the elbow. There were no acceptable PPG waveforms from the elbow in both patients, so these were not used in any of the U-Nets. A single encoder was used for the first to third U-Nets, since the input for these comprised only a single type of waveform (i.e. either blood flow or PPG), whereas two encoders were used for the fourth and fifth U-Nets – one for the blood flow waveform and the other for the PPG waveform (Fig. 3a). Each U-Net had two encoder and decoder layers with two convolutions in each layer and max pooling between layers. Skip connections were also used between the encoder and decoder at each layer to preserve the originality of the feature maps at each layer.

**Fig. 3:**
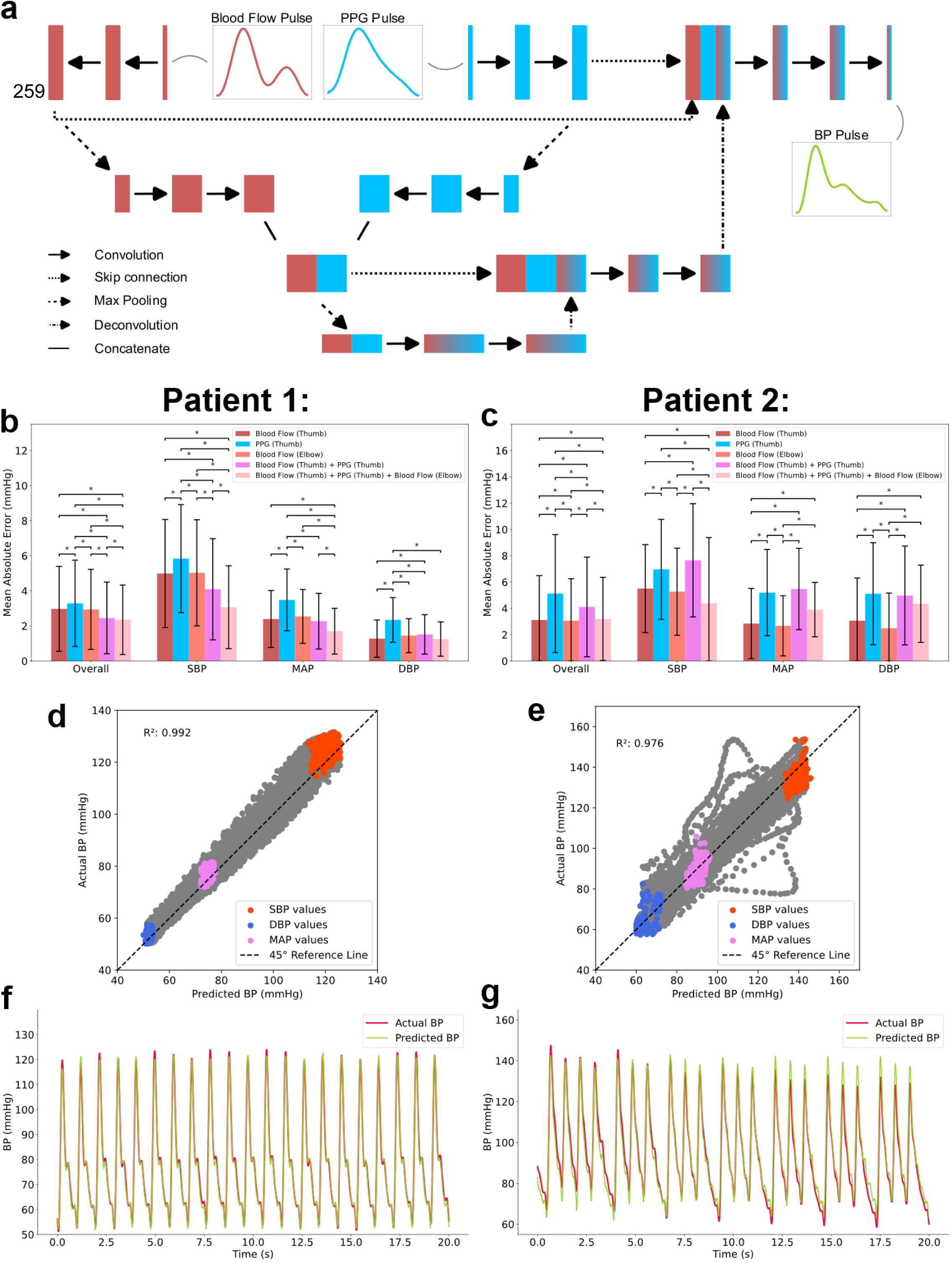
Validation of the DSPF system in the inpatient study. a) Blood flow and/or PPG pulses were used to train U-Nets to predict the BP pulse. Each U-Net had two encoder and two decoder layers with max pooling or deconvolution between layers and skip connections between the encoder and decoder branches at each layer. b-c) Bar charts showing the comparison of MAEs for the overall pulse, SBP, MAP, and DBP in patient 1 (b) and patient 2 (c) across the different U-Nets trained using varying combinations of blood flow and PPG pulses. Error bars represent standard deviation while asterisks represent statistical significance (p < 0.05) between absolute errors from pairs of U-Nets. d-e) Correlation plots between DSPF-predicted BP and BP measured from the A-line in patient 1 (d) and patient 2 (e) show good agreement with high coefficient of determination (R2) values. f-g) Plots of the actual and predicted BP waveforms in patient 1 (f) and patient 2 (g) reveal that the DSPF-predicted BP waveforms possess a morphology that closely matches those obtained via the A-line.

Each U-Net was trained to take in blood flow and/or PPG waveform(s) as the input and produce a BP waveform as the output. The predicted BP waveforms were then compared with the ground truth waveforms from the BP signal. Additionally, the SBP, DBP, and mean arterial pressure (MAP) were determined from the predicted and actual BP waveforms and likewise compared. The overall MAE for each waveform as well as the MAEs for the SBP, MAP, and DBP were calculated to evaluate and compare the performance of the different models (Fig. 3b, c). To determine statistical significance, we similarly performed the Kruskal-Wallis test followed by Dunn’s test with Bonferroni’s correction if required to identify pairs of U-Nets which had a statistically significant difference.

For patient 1, the U-Net trained on all available waveforms (i.e. blood flow and PPG waveforms from the thumb, and blood flow waveforms from the elbow) was the best-performing, achieving the lowest MAEs of 2.35 mmHg for the overall pulse, 3.07 mmHg for SBP, 1.70 mmHg for MAP, and 1.25 mmHg for DBP (Fig. 3b). This is expected as the set of waveforms from the two anatomical sites and two modalities provides a large amount of hemodynamic information, the most possible amount in this study. Using waveforms from both the thumb and elbow allows information on the propagation of pulses through the vasculature that connects them to be captured, providing insight into the mechanical properties of the vessels which strongly influence the BP waveform^49^. The combination of information from the blood flow and PPG waveforms also paints a more comprehensive picture of the underlying hemodynamic processes and therefore a better estimation of the BP waveform. Conversely, the U-Net trained only on PPG waveforms from the thumb was consistently outperformed by all the other U-Nets. In particular, this U-Net produced MAEs which were significantly higher than those attained by the U-Net using the blood flow waveforms from the same location, indicating that blood flow waveforms can be more closely mapped to the BP waveform compared to PPG waveforms.

For patient 2, a similar result was observed between the U-Nets trained on either the blood flow or PPG waveforms from the thumb (Fig. 3c). However, the U-Nets which used a combination of blood flow and PPG waveforms from either the thumb or both the thumb and elbow were often outperformed by a U-Net using only a single type of waveform. The best-performing U-Net for the overall waveform was the one using blood flow waveforms from the elbow which achieved an MAE of 3.06 mmHg. It also had the lowest MAEs for MAP (2.68 mmHg) and DBP (2.49 mmHg), and the second lowest MAE for SBP (5.27 mmHg), suggesting that the elbow can be used as a suitable anatomical site for BP measurement with the DSPF system. The inferior performance of the U-Nets using a combination of blood flow and PPG waveforms can be attributed to the low number of acceptable pulses used to train these networks which we discuss further below.

Correlation plots of the actual and predicted BP values for best-performing U-Nets also indicated good agreement, having R^2^ values of 0.992 for patient 1 (Fig. 3d) and 0.976 for patient 2 (Fig. 3e). SBP, MAP, and DBP values were also in close proximity to the 45° reference line. Although some deviations from the reference line are more noticeable in the plot for patient 2, these arose from a few outlier pulses which were suspected to have occurred due to the patient’s arrhythmia. Despite these isolated pulses, we found that the U-Nets for both patients were able to reconstruct the morphology of the BP waveforms with good accuracy.

This observation was verified in beat-to-beat comparisons of the actual and predicted BP waveforms from each patient (Fig. 3f, g). In the plot for patient 1 (Fig. 3f), we observed that the predicted BP waveforms closely reflected the upstroke (anacrotic) and downstroke (dicrotic) portions of the BP waveform as well as its fiducial points^50^ such as the systolic peak and dicrotic notch. The widths of the predicted waveforms were also well-matched with those of the actual waveforms, and phase alignment was noted. In the plot for patient 2, we also observed that the shape of pulses with irregular pulse intervals was correctly predicted, suggesting that the U-Net can also learn and reconstruct waveforms with abnormal or inconsistent morphologies.

### Further investigation of the DSPF system for BP measurement

To further characterise the performance of the DSPF system for the measurement of BP, we calculated the number of acceptable blood flow and PPG pulses which could be obtained from the recorded signals and subsequently used to train models for BP measurement (Fig. 4a). In the outpatient study, the median number of acceptable blood flow pulses obtained across the 133 recruited patients (over a five-minute recording period) was 286 from the thumb, 226 from the wrist, and 267 from the elbow. On the other hand, the median numbers of acceptable PPG pulses obtained from the thumb, wrist, and elbow were considerably lower at 273, 67, 41, respectively. PPG signals are known to be highly susceptible to noise^51^, a limitation that affects signal quality and consequently the accuracy of BP measurements. Here we find that the blood flow waveforms have a higher fidelity and are less affected by noise-induced distortions.

**Fig. 4:**
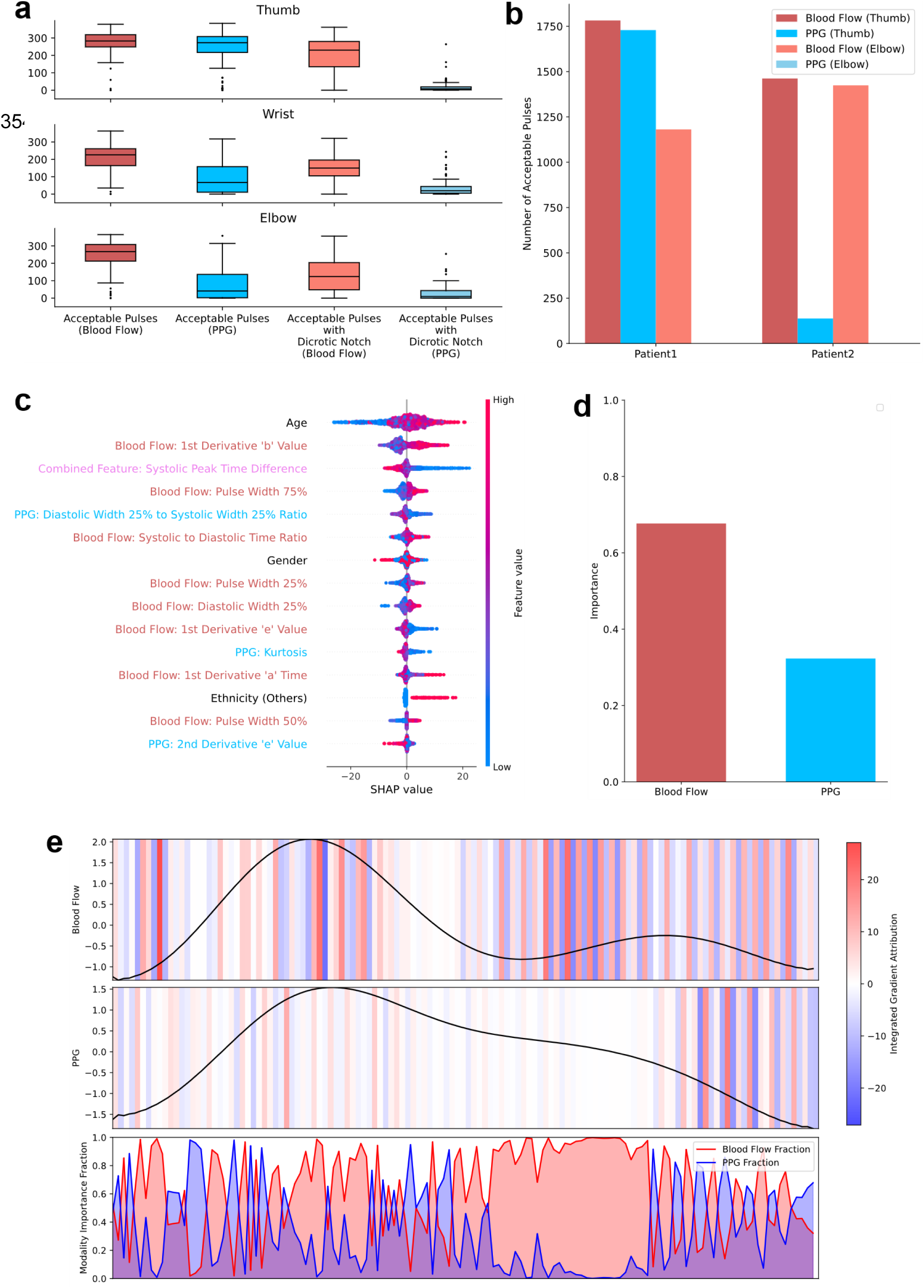
Further characterisation of the DSPF system for BP measurement. a) Box plots of the number of acceptable pulses in the blood flow and PPG signals across all recruited patients in the outpatient study show higher numbers of acceptable blood flow pulses compared to PPG pulses over three anatomical locations. The number of acceptable pulses with a dicrotic notch, an important landmark in the waveform for the extraction of morphological features, is also higher in the blood flow signal. b) Bar charts for the number of acceptable pulses from both the blood flow and PPG signals in the two recruited patients in the inpatient study similarly show that the number of acceptable blood flow pulses is higher than the number of acceptable PPG pulses. Notably, there are no acceptable PPG pulses from the elbow while there is a sizeable yield of acceptable blood flow pulses. c) The SHAP beeswarm plot for the XGBoost model trained on features from both the blood flow and PPG pulses reveal that blood flow and combined features have higher importances in the model’s predictions compared to PPG features. d) Bar chart showing the proportion of importance assigned to blood flow and PPG pulses in the U-Net trained on both modalities for patient 1, as revealed by an integrated gradients analysis. e) Results from the integrated gradients analysis for a representative blood flow and PPG pulse pair corresponding to the same cardiac cycle show the distribution of importance assigned to both modalities over different temporal regions in the waveform. High importance is assigned to the regions around the systolic peak and dicrotic notch of the blood flow waveform.

We also calculated the number of pulses with a dicrotic notch (Fig. 4a), an important landmark for the extraction of many morphological features that contribute to the determination of BP^52^. The median number of acceptable blood flow pulses with a dicrotic notch that could be obtained from a patient was 230 at the thumb, 150 at the wrist, and 124 at the elbow. These numbers similarly exceeded the median numbers of acceptable PPG pulses with a dicrotic notch, which were 8, 19, and 9 at the thumb, wrist, and elbow respectively. The large discrepancies imply that more features and therefore more hemodynamic information can be extracted from the blood flow pulses compared to the PPG pulses.

In the inpatient study, higher numbers of acceptable blood flow pulses compared to PPG pulses were also observed in each patient (Fig. 4b). In patient 1, 1782 acceptable blood flow pulses were obtained from the thumb compared to 1729 acceptable PPG pulses, representing a 3% difference. In patient 2, this difference was much larger at 91%; there were 1462 acceptable blood flow pulses from the thumb but only 138 acceptable PPG pulses. At the elbow, no acceptable PPG pulses could be obtained while there was a high yield (>1000) of blood flow pulses for each patient.

A possible explanation for this observation that is common to both studies is that the effect of random, environmental, and physiological noise is larger in the PPG signal; such noise has a direct and proportional effect on the calculation of the mean pixel value from the laser speckle pattern. In comparison, the calculation of the blood flow signal involves determining the standard deviation of the pixel values then dividing it by the mean pixel value. An identical amount of noise in the blood flow signal would therefore cause disproportionately less distortion, especially at the wrist and elbow where the signal-to-noise ratios are lower due to higher noise levels and greater signal attenuation. The blood flow signal is thus more robust to noise than the PPG signal, particularly at the wrist and elbow, and therefore allows more pulses of acceptable signal quality to be obtained for the training of BP models, as well as the calculation of BP.

The shortage of acceptable PPG pulses from the thumb signal of patient 2 in the inpatient study is likely to be the cause of the lower accuracies of the U-Nets which use a combination of blood flow and PPG pulses to predict the BP pulse. To ensure proper training of these U-Nets, we only used blood flow and PPG waveforms corresponding to the same heartbeat as training samples. The number of samples was therefore limited by the number of acceptable PPG pulses which was severely lower than the number of acceptable blood flow pulses.

Aside from the higher number of acceptable pulses, the blood flow waveforms provided information that was more beneficial and important for the estimation of BP compared to the PPG waveforms. In the outpatient study, an analysis of the Shapley additive explanation (SHAP) values for the XGBoost model trained on blood flow, PPG, and combined features from the thumb showed that the 15 most important features consisted of eight blood flow features, one combined feature, three PPG features, and three demographic features (Fig. 4c). The top feature was age, with which BP is known to increase with^53^. Following age, the ‘b’ value in the first derivative of the blood flow waveform was the second most important feature. It represents the greatest deceleration of blood flow during systole, or the early systolic negative wave^54^ which is influenced by the distensibility of the arteries^55^, a key determinant of BP^56^. The third most important feature was the time difference between the systolic peaks of the blood flow and PPG waveforms, which is similarly related to the viscoelasticity of the arteries and consequently BP. Other top features included different widths within the blood flow and PPG pulses, as well as other features from the first derivative of the blood flow waveform and second derivative of the PPG waveform.

Age accounted for 17% of the model’s decision across all predictions, while the ‘b’ value of the first derivative of the blood flow waveform accounted for 12%, and the systolic peak time difference accounted for 8%. Collectively, the blood flow features contributed to 43% of the model’s decision, while combined features had a contribution of 10%, PPG features a contribution of 22%, and demographic features a contribution of 25%. This finding shows the higher importance of blood flow features to BP measurement compared to PPG features, and therefore the ability of the DSPF system to augment the performance of PPG-based BP models with the addition of these as well as combined features.

The blood flow waveform also had a higher importance compared to the PPG waveform in the inpatient study. Through an integrated gradients analysis of the U-Net that used both the blood flow and PPG pulses from patient 1 which we performed to identify which modality was a more important contributor to the predicted BP waveforms, it was found that the blood flow pulses had a higher importance proportion of 0.677 while the PPG pulses had a lower importance fraction of 0.323 (Fig. 4d). This is similar to what was revealed by the SHAP analysis, where the contribution of the blood flow waveform to the model’s decision was approximately twice that of the PPG waveform.

As can be seen from an example of the distribution of importance between a representative blood flow and PPG waveform pair (Fig. 4e), the blood flow waveform dominates the PPG waveform in importance over many temporal regions across the duration of the pulse. This is particularly so around the early upstroke period, as well as the periods surrounding the systolic peak and dicrotic notch. These regions within the blood flow waveform thus have a higher explanatory power than their PPG counterparts, and may provide finer detail to augment the construction of the BP pulse with better conformity to the shape of ground truth pulse with its morphological intricacies. The high importance allocated to the region around the dicrotic notch also indicates the importance of having this fiducial point in the waveform, which has a higher availability in the blood flow waveform as mentioned earlier.

## Discussion

Across both the outpatient and inpatient studies, we find that the DSPF system is capable of achieving high accuracies for the measurement of BP. In the outpatient study, a low MAE of < 5 mmHg was reached with the waveforms obtained from the thumb. The mean error was also < 5 mmHg while the standard deviation of the errors was < 8 mmHg, satisfying criterion 1 of the AAMI/ESH/ISO standard. Importantly, this was reached while satisfying other criteria of the standard which concerned the number of recruited patients and the distribution of reference SBP and DBP values presented by the patients. An ‘A’ grade rating was also achieved based on the BHS’s guidelines.

In the inpatient study, the DSPF system could similarly achieve low MAEs of < 3.5 mmHg for the overall pulse, MAP, and DBP, and < 6 mmHg for SBP in the two recruited patients. Interestingly, the best-performing model in one patient (patient 2) was the one trained on blood flow pulses from the elbow. In the other patient (patient 1) this same type of model did not have a significantly different performance compared to that trained on blood flow pulses from the thumb. This highlights the feasibility of using the elbow as an anatomical site for beat-to-beat BP measurement using the DSPF system, which may be useful especially if a patient’s wrist or thumb is required for the placement of other equipment or accessories (e.g. intravenous drip, identification tag, pulse oximeter). On the contrary, PPG systems would fail at the elbow because of the low signal-to-noise ratio.

We also found that the DSPF system improves the performance of PPG-only methods for BP measurement through the addition of the blood flow rate signal. The benefit of this is two-fold; the blood flow rate signal is more robust to noise and has a higher signal quality, which enables more pulses of acceptable quality to be extracted from it. This consequently improves the training of BP models and their predictions. Separately, the blood flow waveform was a more important contributor to the predictions of the BP models for the outpatient and inpatient studies compared to the PPG waveform, which suggests that the hemodynamic information contained within it is more pertinent to the measurement of BP.

Overall, we find the DSPF system to be a viable alternative to current BP measurement devices and techniques. In outpatient settings, BP can be obtained without a cuff using DSPF sensors fitted into an oximeter-like thumb clip. With this setup, clinicians who take BP measurements from patients would no longer need to fit a cuff around patients’ arms and ensure that it is positioned appropriately but instead benefit from the convenience of simply opening and closing a thumb clip. The DSPF system also requires a much shorter time to calculate a BP reading compared to cuff-based methods, and thus has the potential to improve the output and efficiency of clinicians in addressing patients’ needs. In addition, using DSPF enables BP to be measured continuously, which allows a second or repeat set of BP readings to be obtained. This may provide a more accurate assessment of BP as the second reading is preferred to mitigate the effects of any white-coat hypertension in the clinic or even in out-of-clinic settings^57^.

In the inpatient setting, using DSPF sensors may also reduce or eliminate the need for invasive A-lines which pose the risk of infection and cause discomfort to patients. Future efforts to miniaturise the DSPF system may also allow for its inclusion in current consumer and medical-grade wearables, which could enhance the measurement of BP that presently involves PPG instrumentation only. We hence foresee that the DSPF system can improve the measurement of BP in both outpatient, inpatient, and remote settings, whether from a practical perspective by overcoming the limitations of cuff-based and invasive methods, or from a performance perspective by improving the accuracy of PPG-based methods.

## Methods

### A: DSPF Instrumentation and Operation

The DSPF system consisted of two to three channels of DSPF sensors; three channels were used for the outpatient study whereas two were used for the inpatient study. Each channel was made up of an 850 nm laser diode operated at ≤ 5 mW, a source-detector pair of 200 µm multimode optic fibres (ThorLabs), and a CCD (ThorLabs). Light emitted from the laser was passed through the source fibre to illuminate the skin, while the detector fibre captured backscattered photons and transmitted them to the CCD which recorded a laser speckle pattern. The distance between the two fibres (source-detector separation) was 15 mm.

The system was operated and controlled by a custom LabVIEW program and graphical user interface. Data from all channels was simultaneously recorded at a sampling rate of 300 Hz for the outpatient study and 330 Hz for the inpatient study, a parameter determined by the CCD’s frame rate setting. Upon commencement of a recording, 6000 image frames which each displayed a laser speckle pattern were averaged to produce a background intensity profile. This measured profile was used to normalise and correct the intensity distribution of subsequent speckle patterns from the tip of the detector fibre which would be distorted because of a non-uniform beam shape^19^.

Statistical measures of the corrected speckle pattern at each frame were then determined to extract the instantaneous blood flow rate and blood volume. Speckle contrast, defined as the ratio of the mean pixel intensity to the standard deviation of pixel intensities, was calculated and the inverse of its squared value was taken as a measure of blood flow rate^19^. Meanwhile, blood volume was calculated as the mean pixel intensity of the speckle pattern. Should readers desire more details on the theory of operation, these can be found in earlier studies^19,58^.

### B: Experimental Protocol (Outpatient Study)

133 patients were recruited at a primary care facility (Eunos Polyclinic, Singapore Health Services, Singapore) for the outpatient study. Each subject was at least 21 years old, without cognitive impairment, without upper limb deformity or pain, not diagnosed with atrial fibrillation, and not pregnant. The study procedure was explained to the satisfaction of each participant, and all participants signed an informed consent form prior to the commencement of the study. In addition, each participant completed a data collection form where they provided information about their age, gender, ethnicity, and prevailing medical conditions. Ethical approval for the study was granted by the SingHealth Centralised Institutional Review Board (ref. 2023/2501).

After the study procedure was explained and informed consent was obtained, each patient was asked to sit on a chair and adopt a comfortable position. His/her arm was then placed on a cushion atop an adjacent desk at the level of the heart. A deflated BP cuff connected to a US FDA-approved sphygmomanometer (HEM 7200, Omron) was then placed on each patient’s upper arm. The cuff was wrapped around the left arm for all except five patients who requested for the measurements to be obtained from the right arm due to previous injections or surgeries which had been performed on their left arm. Care was taken to ensure that the cuff was properly positioned according to the American Heart Association’s guidelines^59^; 2 to 3 cm above the antecubital fossa, at an appropriate level of tightness, and with the indicator arrow aligned with the brachial artery.

Three sets of DSPF sensors were then placed on the patient’s arm at locations distal to the BP cuff – one set over the brachial artery at the region of the antecubital fossa, one set over the radial artery at the wrist, and one set at the thumb. The brachial and radial arteries were identified by palpation. The DSPF sensors over the brachial and radial arteries were secured via wrapping elastic Velcro straps around the circumference of the patient’s arm, while the sensor at the thumb was fitted into a pulse oximeter-like thumb clip into which the patient inserted his/her thumb.

Once the patient was comfortable and ready for DSPF signals to be recorded, background correction was applied and the DSPF blood flow and blood volume signals were synchronously recorded at all three locations for 5 min. Thereafter, the cuff was inflated and SBP and DBP measurements were obtained using the sphygmomanometer. Patients were advised to relax and not talk throughout the course of the measurements. After the DSPF signals and BP measurements had been recorded, the DSPF sensors and BP cuff were removed from the patient’s arm.

### C: Experimental Protocol (Inpatient Study)

Two patients were recruited for the inpatient study at a specialised cardiology centre (National Heart Centre, Singapore). Each patient was warded and required an A-line for continuous BP monitoring. Both patients were aged between 21 and 90 years old, not pregnant or breastfeeding, and not diagnosed with severe heart failure, malignancy, end stage renal failure, or significant neuromuscular disease. The A-line was inserted into the radial artery, while DSPF sensors were placed over the brachial artery in the antecubital fossa region and the thumb for each patient. Like in the outpatient study, elastic Velcro straps were wrapped around the patient’s arm to secure the DSPF sensors over the brachial artery and a thumb clip was used to house the DSPF sensors which were placed over the thumb.

Both patients were similarly briefed on the experimental procedure and allowed to ask any questions. Once this had been appropriately completed, each patient signed an informed consent form. The ethical approval for this study was granted by the SingHealth Centralised Institutional Review Board (ref. 2021/2178).

### D: Data Processing

A total of six signals were recorded from each patient in the outpatient study (blood flow and blood volume signals from three locations) while four signals were recorded from each patient in the inpatient study (blood flow and blood volume signals from two locations). After the signals were obtained, each was bandpass-filtered with a 5^th^-order Chebyshev type II bandpass filter from 0.5 to 4 Hz to remove baseline drift and high frequency noise. The A-line BP signal obtained from the patients in the inpatient study was sampled at 125 Hz and lowpass-filtered using a 5^th^-order Chebyshev type II filter with a cutoff frequency of 8 Hz. All processing was done in Python.

In the outpatient study, the DSPF signals were split into non-overlapping windows of 15 s, resulting in 20 windows per signal for each subject. This was done to avoid excessive temporal averaging of the morphological features from the pulses as well as to obtain more samples for subsequent training and testing of machine learning models. Individual pulses were then identified from each window by detecting the troughs representing their onset and end. The troughs were detected by inverting the signals and applying the SciPy peak detection algorithm.

The cluster analysis method proposed by Waugh et al.^60^ was then adapted and applied to remove motion artefacts and poor-quality pulses. The method is based on the assumption that true, clean pulses from an individual have a morphological shape that is generally consistent across cardiac cycles. This shape comprises a period of increasing blood flow or blood volume up to a main peak, followed by a period of decreasing blood flow or blood volume which is physiologically consistent with the processes of systole and diastole. Notches and secondary peaks may also be present in the period of descending blood flow or blood volume. The method also assumes that motion artefacts or any other form of noise which may corrupt pulses in a signal are random and irregular. As such, pulses in the recorded signal which do not have a similar shape to a minimum number of other pulses are likely to be corrupted by real-world artefacts. These pulses are identified and subsequently discarded. Conversely, pulses which are similar in shape to others are considered to have the correct morphology.

In each window, every identified pulse was min-max normalised in amplitude and time to the range [0,1]. Each normalised pulse was resampled to 256 samples, then compared to every other pulse in the window. For each pulse, pulse pairs were formed between the pulse and every other pulse. The RMSE for each pulse pair was computed, and if it was < 0.1 the other pulse was stored in an array which represented the cluster of similar pulses. If the number of other pulses in the array was > 20% of the total number of pulses in the window after the RMSE for all pulse pairs had been calculated, the pulse was considered to be of acceptable quality and kept for subsequent analysis. Otherwise, the pulse was deemed sufficiently dissimilar and therefore discarded.

The similar pulses within each window were retrieved from the filtered signal in their non-normalised form and concatenated to form a sequence. Z-score normalisation was then applied to the sequence, and several pulse acceptability criteria, such as having a minimum of one detected peak and having the number of detected peaks exceed the number of intermediate notches by one were applied. This was done to ensure that morphological features could be appropriately extracted from the pulses.

Features were then extracted from each of the similar and accepted blood flow and blood volume pulses. The features (described in Supplementary Tables 1, 2) can be classified into amplitude-based features, time-based features, features based on areas under the curve, and features that are calculated from linear transformations of one or more features. Features from the first derivative of the blood flow waveform, as well as the first and second derivatives of the blood volume waveform were also calculated. If the blood flow waveform and blood volume waveform from the same pulse both passed the similarity check and were considered as acceptable, combined features involving both these waveforms were also calculated (Supplementary Tables 3).

For the inpatient study, individual pulses from the BP and DSPF signals from each patient were similarly identified via inverting the signal and applying the SciPy peak detection algorithm to locate the troughs which demarcate the onset and termination of each pulse. The pulses in the BP signal were then divided into training, validation, and test sets; the first 60% of pulses were assigned to the training set, the next 20% to the validation set, and the final 20% to the test set. Each pulse from the four DSPF signals was registered with its corresponding pulse in the BP signal.

The pulses identified from the DSPF signals were also assessed using the cluster analysis method applied in the outpatient study. However, instead of windowing the signal and checking for pulse similarity among the pulses within each window, the pulses in the training set were compared with each other. For the validation and test sets, each pulse was compared with the set of similar pulses in the training set. This form of implementation was chosen with the view that it can be applied for real-time BP estimation as new pulses are recorded.

Like in the outpatient study, similar pulses were retained while dissimilar ones were discarded. The retained pulses were then normalised and resampled to 256 samples. This was done to ensure a uniform pulse length that would facilitate the subsequent training of neural networks for BP prediction. Pulse acceptability criteria were also applied to ensure that the morphology of the pulses were physiologically feasible.

### E: BP Prediction

For the outpatient study, median values of the morphological and statistical features extracted from each acceptable pulse were calculated from all acceptable pulses in each window. These, together with demographic features, were used to train separate XGBoost regression models for the prediction of SBP and DBP. Numeric features were normalised with Z-score normalisation and categorical features (i.e. gender, ethnicity) were encoded with dummy variables.

An ablation study was performed, where different feature sets were used to train separate XGBoost models. Three feature sets were used from the signals at each anatomical location; the first comprising blood flow features, the second comprising PPG features, and the third comprising both blood flow and PPG features. Each XGBoost model was trained using five-fold cross-validation with 400 estimators and a maximum of 30 features. To evaluate the models, the MAE, mean error, and standard deviation of errors was calculated between the predicted SBP or DBP values and the actual values measured by the sphygmomanometer.

For the inpatient study, an ablation study was also performed where separate U-Net models were trained to reconstruct BP waveforms. A total of five U-Nets were trained for each patient using the normalised and resampled pulses from individual DSPF signals and a combination of them. The first three were trained on pulses from individual DSPF signals – one was trained on pulses in the blood flow signal from the thumb, the second on pulses in the PPG signal from the thumb, and the third on pulses in the blood flow signal from the elbow. No model was trained using PPG pulses from the elbow as there were no acceptable pulses which could be used. The fourth U-Net used a combination of blood flow and PPG pulses from the thumb while the fifth used the blood flow and PPG pulses from the thumb as well as the blood flow pulses from the elbow.

A 1-dimensional (1D) U-Net with a single encoder branch was trained when pulses of a single type of signal were used (i.e. blood flow pulses or PPG pulses), while a U-Net with two encoder branches was trained when both blood flow and PPG pulses were used. Aside from these differences, all models were trained with the Adam optimiser across 100 epochs. The BP pulses were normalised with Z-score normalisation prior to training such that the model could learn the BP waveform morphology, and the training data was input into the model in randomised batches of size 32. At each epoch, the loss was calculated as a weighted sum of the mean squared error (MSE) of the actual pulses and the MSE of the pulses’ first derivative. The predicted BP waveforms were then scaled back to their original range and the mean as well as standard deviation of the absolute errors for the overall waveform, SBP, MAP, and DBP were calculated.

## Data Availability

The patient data from the present study is not publicly available.

## Acknowledgements

The authors thank the clinical research coordinators and assistants who helped with the recruitment and administration of the study. We also acknowledge scottdejonge for the use of the SVG icon in Fig. 2a from SVG Repo (https://www.svgrepo.com/svg/366768/male) which is licensed under the MIT License.

## Author Contributions

1. T. W. J. C., V. T. T. C., C. J. N., and R. B. conceptualised the work; T. W. J. C., A. K., V. T. T. C., C. J. N., and R. B. designed, developed, and refined the methodology; T. W. J. C., A. A. M. Y., and A. K. conducted the experiments and investigation; T. W. J. C. and A. A. M. Y. performed the formal analysis; T. W. J. C., C. J. N., and R. B. contributed to the writing, reviewing, and editing of the manuscript; T. W. J. C. and R. B. prepared the figures; V. T. T. C., C. J. N., M. O., and R. B. provided the necessary resources, funding, project administration, and supervision.

---

**Supplementary Table 1:** Features extracted from the blood flow waveform.

| Waveform Type | Feature | Description |
| --- | --- | --- |
| Blood Flow | Systolic Peak Amplitude | Difference between the systolic peak value and the value at pulse onset. |
|  | Second Peak Amplitude | Difference between the value of the waveform's second peak and the value at pulse onset. |
|  | Pulse Interval | Duration of the pulse from onset to termination. |
|  | Peak Systolic Speed | Maximum value of the waveform. |
|  | End Diastolic Speed | Value of the waveform at termination. |
|  | Minimum Speed | Minimum value of the waveform. |
|  | Resistive Index | Difference between the peak systolic speed and end diastolic speed, normalised by the peak systolic speed. |
|  | Pulsatility Index | Difference between the peak systolic speed and end diastolic speed, normalised by the mean pulse speed. |
|  | Mean | Mean value of the waveform across time. |
|  | Standard Deviation | Standard deviation of the waveform values across time. |
|  | Skewness | Skewness coefficient of the waveform values across time. |
|  | Kurtosis | Kurtosis coefficient of the waveform values across time. |
|  | Systolic to Diastolic Ratio | Ratio of the peak systolic speed to end diastolic speed. |
|  | Systolic to Diastolic Time Ratio | Ratio of the time from pulse onset to systolic peak and the time from systolic peak to pulse termination. |
|  | Acceleration Index | Ratio of the difference between the peak systolic speed and pulse onset speed to the time between pulse onset and the systolic peak. |
|  | Rise Time | Time from the pulse onset to the systolic peak. |
|  | Decay Time | Time from the systolic peak to pulse termination. |
|  | Augmentation Index | Ratio of the systolic peak amplitude to the second peak amplitude. |
|  | Systolic Width 25% | Time difference between the systolic peak and the point on the rising limb of the waveform having an amplitude equal to 25% of the systolic peak amplitude. |
|  | Systolic Width 50% | Time difference between the systolic peak and the point on the rising limb of the waveform having an amplitude equal to 50% of the systolic peak amplitude. |
|  | Systolic Width 75% | Time difference between the systolic peak and the point on the rising limb of the waveform having an amplitude equal to 75% of the systolic peak amplitude. |
|  | Diastolic Width 25% | Time difference between the systolic peak and the point on the descending limb of the waveform having an amplitude equal to 25% of the systolic peak amplitude. |
|  | Diastolic Width 50% | Time difference between the systolic peak and the point on the descending limb of the waveform having an amplitude equal to 50% of the systolic peak amplitude. |
|  | Diastolic Width 75% | Time difference between the systolic peak and the point on the descending limb of the waveform having an amplitude equal to 75% of the systolic peak amplitude. |
| Blood Flow 1<sup>st</sup> Derivative | 'a' value | Value of the 'a' point in the 1<sup>st</sup> derivative of the blood flow waveform. The 'a' point is the first peak in the 1<sup>st</sup> derivative of the blood flow waveform. |
|  | 'b' value | Value of the 'b' point in the 1<sup>st</sup> derivative of the blood flow waveform. The 'b' point is the first trough in the 1<sup>st</sup> derivative of the blood flow waveform. |
|  | 'c' value | Value of the 'c' point in the 1<sup>st</sup> derivative of the blood flow waveform. The 'c' point is the local maximum in the 1<sup>st</sup> derivative of the blood flow waveform that occurs during the period between the systolic peak and aortic notch in the blood flow waveform. |
|  | 'd' value | Value of the 'd' point in the 1<sup>st</sup> derivative of the blood flow waveform. The 'd' point is the local minimum in the 1<sup>st</sup> derivative of the blood flow waveform that occurs after the 'c' point and during the period between the systolic peak and aortic notch in the blood flow waveform. |
|  | 'e' value | Value of the 'e' point in the 1<sup>st</sup> derivative of the blood flow waveform. The 'e' point is the local maximum in the 1<sup>st</sup> derivative of the blood flow waveform that occurs during the period between the aortic notch and second peak in the blood flow waveform. |
|  | 'a' time | Time from the onset to the 'a' point in the 1<sup>st</sup> derivative of the blood flow waveform. |
|  | 'b' time | Time from the onset to the 'b' point in the 1<sup>st</sup> derivative of the blood flow waveform. |
|  | 'c' time | Time from the onset to the 'c' point in the 1<sup>st</sup> derivative of the blood flow waveform. |
|  | 'd' time | Time from the onset to the 'd' point in the 1<sup>st</sup> derivative of the blood flow waveform. |
|  | 'e' time | Time from the onset to the 'e' point in the 1<sup>st</sup> derivative of the blood flow waveform. |
|  | 'b' to 'c' time | Time between the 'b' and 'c' points in the 1<sup>st</sup> derivative of the blood flow waveform. |
|  | 'b' to 'd' time | Time between the 'b' and 'd' points in the 1<sup>st</sup> derivative of the blood flow waveform. |
|  | 'b' to 'a' ratio | Ratio of the 'b' and 'a' values in the 1<sup>st</sup> derivative of the blood flow waveform. |
|  | 'c' to 'a' ratio | Ratio of the 'c' and 'a' values in the 1<sup>st</sup> derivative of the blood flow waveform. |
|  | 'd' to 'a' ratio | Ratio of the 'd' and 'a' values in the 1<sup>st</sup> derivative of the blood flow waveform. |
|  | 'e' to 'a' ratio | Ratio of the 'e' and 'a' values in the 1<sup>st</sup> derivative of the blood flow waveform. |
|  | 'b' to 'c' slope | Gradient between the 'b' and 'c' points in the 1<sup>st</sup> derivative of the blood flow waveform. |
|  | 'b' to 'd' slope | Gradient between the 'b' and 'd' points in the 1<sup>st</sup> derivative of the blood flow waveform. |
|  | Aging Index | 'b' value minus 'c' value minus 'd' value minus 'e' value, divided by the 'a' value. |
|  | Modified Aging Index | 'b' value minus 'c' value minus 'd' value, divided by the 'a' value. |
|  | Informal Aging Index | 'b' value minus 'e' value, divided by the 'a' value. |

**Supplementary Table 2:** Features extracted from the PPG waveform.

| Waveform Type | Feature | Description |
| --- | --- | --- |
| PPG | Systolic Peak Amplitude | Difference between the systolic peak value and the value at pulse onset. |
|  | Systolic Peak Time | Time from the pulse onset to the systolic peak. |
|  | Pulse Interval | Duration of the pulse from onset to termination. |
|  | Systolic Peak Amplitude over Systolic Time | Ratio between the systolic peak amplitude and systolic peak time. |
|  | Diastolic Peak Amplitude | Difference between the diastolic peak value and the value at pulse onset. |
|  | Diastolic Peak Time | Time from the pulse onset to the diastolic peak. |
|  | Dicrotic Notch Amplitude | Difference between the dicrotic notch value and the value at pulse onset. |
|  | Dicrotic Notch Time | Time from the pulse onset to the dicrotic notch. |
|  | Time Delta | Time between the systolic and diastolic peaks. |
|  | Systolic Peak Time over Dicrotic Notch Time | Ratio of the systolic peak time to the dicrotic notch time. |
|  | Mean | Mean value of the waveform across time. |
|  | Standard Deviation | Standard deviation of the waveform values across time. |
|  | Skewness | Skewness coefficient of the waveform values across time. |
|  | Kurtosis | Kurtosis coefficient of the waveform values across time. |
|  | Augmentation Index | Ratio of the systolic peak amplitude to the diastolic peak amplitude. |
|  | Alternative Augmentation Index | Difference between the systolic peak amplitude and diastolic peak amplitude, divided by the diastolic peak amplitude. |
|  | Dicrotic Notch Amplitude over Systolic Peak Amplitude | Ratio of the dicrotic notch amplitude to the systolic peak amplitude. |
|  | Dicrotic Notch Time over Dicrotic Notch Amplitude | Ratio of the dicrotic notch time to the dicrotic notch amplitude. |
|  | Diastolic Peak Time over Diastolic Peak Amplitude | Ratio of the diastolic peak time to the diastolic peak amplitude. |
|  | Systolic Peak Amplitude over Diastolic Time | Ratio of the systolic peak amplitude to the time between the systolic peak and pulse termination. |
|  | Systolic Width 25% | Time difference between the systolic peak and the point on the rising limb of the waveform having an amplitude equal to 25% of the systolic peak amplitude. |
|  | Systolic Width 50% | Time difference between the systolic peak and the point on the rising limb of the waveform having an amplitude equal to 50% of the systolic peak amplitude. |
|  | Systolic Width 75% | Time difference between the systolic peak and the point on the rising limb of the waveform having an amplitude equal to 75% of the systolic peak amplitude. |
|  | Diastolic Width 25% | Time difference between the systolic peak and the point on the descending limb of the waveform having an amplitude equal to 25% of the systolic peak amplitude. |
|  | Diastolic Width 50% | Time difference between the systolic peak and the point on the descending limb of the waveform having an amplitude equal to 50% of the systolic peak amplitude. |
|  | Diastolic Width 75% | Time difference between the systolic peak and the point on the descending limb of the waveform having an amplitude equal to 75% of the systolic peak amplitude. |
|  | Pulse Width 25% | Time difference between the point on the rising limb of the waveform and the point on the descending limb of the waveform having amplitudes equal to 25% of the systolic peak amplitude. |
|  | Pulse Width 50% | Time difference between the point on the rising limb of the waveform and the point on the descending limb of the waveform having amplitudes equal to 50% of the systolic peak amplitude. |
|  | Pulse Width 75% | Time difference between the point on the rising limb of the waveform and the point on the descending limb of the waveform having amplitudes equal to 75% of the systolic peak amplitude. |
|  | Diastolic Width over Systolic Width 25% | Ratio of diastolic width 25% to systolic width 25%. |
|  | Diastolic Width over Systolic Width 50% | Ratio of diastolic width 50% to systolic width 50%. |
|  | Diastolic Width over Systolic Width 75% | Ratio of diastolic width 75% to systolic width 75%. |
|  | Area Under Curve | Area under the waveform curve from pulse onset to pulse termination. |
|  | Systolic Area | Area under the waveform curve from pulse onset to the dicrotic notch. |
|  | Diastolic Area | Area under the waveform curve from the dicrotic notch to pulse termination. |
|  | Inflection Point Area | Ratio of the diastolic area to the systolic area. |
| PPG 1<sup>st</sup> Derivative | 'w' value | Value of the 'w' point in the 1<sup>st</sup> derivative of the PPG waveform. The 'w' point is the first peak in the 1<sup>st</sup> derivative of the PPG waveform. |
|  | 'y' value | Value of the 'y' point in the 1<sup>st</sup> derivative of the PPG waveform. The 'y' point is the first trough in the 1<sup>st</sup> derivative of the PPG waveform. |
|  | 'z' value | Value of the 'z' point in the 1<sup>st</sup> derivative of the PPG waveform. The 'z' point is the second peak in the 1<sup>st</sup> derivative of the PPG waveform. |
|  | 'w' time | Time from the onset to the 'w' point in the 1<sup>st</sup> derivative of the PPG waveform. |
|  | 'y' time | Time from the onset to the 'y' point in the 1<sup>st</sup> derivative of the PPG waveform. |
|  | 'z' time | Time from the onset to the 'z' point in the 1<sup>st</sup> derivative of the PPG waveform. |
|  | Maximum Slope over Systolic Peak Amplitude | Ratio of the 'w' value in the 1<sup>st</sup> derivative of the PPG waveform to the systolic peak amplitude in the PPG waveform. |
|  | Systolic Mean | The mean value of the waveform between the pulse onset and systolic peak. |
|  | Systolic Variance | The variance of the waveform values between the pulse onset and systolic peak. |
|  | Diastolic Mean | The mean value of the waveform between the systolic peak and pulse termination. |
|  | Diastolic Variance | The variance of the waveform values between the systolic peak and pulse termination. |
| PPG 2<sup>nd</sup><br>Derivative | 'a' value | Value of the 'a' point in the 2<sup>nd</sup> derivative of the PPG waveform. The 'a' point is the first peak in the 2<sup>nd</sup> derivative of the PPG waveform. |
|  | 'b' value | Value of the 'b' point in the 2<sup>nd</sup> derivative of the PPG waveform. The 'b' point is the first trough in the 2<sup>nd</sup> derivative of the PPG waveform. |
|  | 'c' value | Value of the 'c' point in the 2<sup>nd</sup> derivative of the PPG waveform. The 'c' point is the local maximum in the 2<sup>nd</sup> derivative of the PPG waveform that occurs during the period between the first peak and first notch in the 1<sup>st</sup> derivative of the PPG waveform. |
|  | 'd' value | Value of the 'd' point in the 2<sup>nd</sup> derivative of the PPG waveform. The 'd' point is the local minimum in the 2<sup>nd</sup> derivative of the PPG waveform that occurs after the 'c' point and during the period between the first peak and first notch in the 1<sup>st</sup> derivative of the PPG waveform. |
|  | 'e' value | Value of the 'e' point in the 2<sup>nd</sup> derivative of the PPG waveform. The 'e' point is the local maximum in the 2<sup>nd</sup> derivative of the PPG waveform that occurs during the period between the first notch and second peak in the 1<sup>st</sup> derivative of the PPG waveform. |
|  | 'a' time | Time from the onset to the 'a' point in the 2<sup>nd</sup> derivative of the PPG waveform. |
|  | 'b' time | Time from the onset to the 'b' point in the 2<sup>nd</sup> derivative of the PPG waveform. |
|  | 'c' time | Time from the onset to the 'c' point in the 2<sup>nd</sup> derivative of the PPG waveform. |
|  | 'd' time | Time from the onset to the 'd' point in the 2<sup>nd</sup> derivative of the PPG waveform. |
|  | 'e' time | Time from the onset to the 'e' point in the 2<sup>nd</sup> derivative of the PPG waveform. |
|  | 'b' to 'c' time | Time between the 'b' and 'c' points in the 2<sup>nd</sup> derivative of the PPG waveform. |
|  | 'b' to 'd' time | Time between the 'b' and 'd' points in the 2<sup>nd</sup> derivative of the PPG waveform. |
|  | 'b' to 'a' ratio | Ratio of the 'b' and 'a' values in the 2<sup>nd</sup> derivative of the PPG waveform. |
|  | 'c' to 'a' ratio | Ratio of the 'c' and 'a' values in the 2<sup>nd</sup> derivative of the PPG waveform. |
|  | 'd' to 'a' ratio | Ratio of the 'd' and 'a' values in the 2<sup>nd</sup> derivative of the PPG waveform. |
|  | 'e' to 'a' ratio | Ratio of the 'e' and 'a' values in the 2<sup>nd</sup> derivative of the PPG waveform. |
|  | 'b' to 'c' slope | Gradient between the 'b' and 'c' points in the 2<sup>nd</sup> derivative of the PPG waveform. |
|  | 'b' to 'd' slope | Gradient between the 'b' and 'd' points in the 2<sup>nd</sup> derivative of the PPG waveform. |
|  | Aging Index | 'b' value minus 'c' value minus 'd' value minus 'e' value, divided by the 'a' value. |
|  | Modified Aging Index | 'b' value minus 'c' value minus 'd' value, divided by the 'a' value. |
|  | Informal Aging Index | 'b' value minus 'e' value, divided by the 'a' value. |

**Supplementary Table 3:** Combined features extracted from the blood flow and PPG waveforms.

| Feature | Description |
| --- | --- |
| Systolic Peak Time Delta | Time between the systolic peaks of the blood flow and PPG waveforms. |
| Second Peak Time Delta | Time between the second peak of the blood flow waveform and the diastolic peak of the PPG waveform. |
| Systolic Peak Amplitude Ratio | Ratio of the systolic peak amplitude of the blood flow waveform to that of the PPG waveform. |
| Second Peak Amplitude Ratio | Ratio of the second peak amplitude of the blood flow waveform to the diastolic peak amplitude of the PPG waveform. |
| Notch Amplitude Ratio | Ratio of the notch amplitude of the blood flow waveform to that of the PPG waveform. |
| Systolic Gradients Product | Product of the acceleration index of the blood flow waveform and the systolic peak amplitude over systolic peak time of the PPG waveform. |
| Augmentation Index Ratio | Ratio of the augmentation indexes of the blood flow and PPG waveforms. |
| Systolic Width 25% Difference | Difference between the systolic width 25% of the blood flow waveform and that of the PPG waveform. |
| Systolic Width 50% Difference | Difference between the systolic width 50% of the blood flow waveform and that of the PPG waveform. |
| Systolic Width 75% Difference | Difference between the systolic width 75% of the blood flow waveform and that of the PPG waveform. |
| Diastolic Width 25% Difference | Difference between the diastolic width 25% of the blood flow waveform and that of the PPG waveform. |
| Diastolic Width 50% Difference | Difference between the diastolic width 50% of the blood flow waveform and that of the PPG waveform. |
| Diastolic Width 75% Difference | Difference between the diastolic width 75% of the blood flow waveform and that of the PPG waveform. |
| Pulse Width 25% Difference | Difference between the pulse width 25% of the blood flow waveform and that of the PPG waveform. |
| Pulse Width 50% Difference | Difference between the pulse width 50% of the blood flow waveform and that of the PPG waveform. |
| Pulse Width 75% Difference | Difference between the pulse width 75% of the blood flow waveform and that of the PPG waveform. |

